# COVID-19 Knowledge, Perceptions, and Behaviors: A Multinational Comparative Study among Bangladesh, Pakistan, and the United States

**DOI:** 10.1101/2025.09.21.25336303

**Authors:** Md. Abdur Rahman, Saiful Islam, Md. Shakibul Hasan, Md. Mostafizur Rahman, Md. Abdul Khalek, Md. Palash Bin Faruque, Mst. Shabrina Afroz

**Affiliations:** Department of Sociology, Noakhali Science and Technology University, Noakhali-3814, Bangladesh; Data Mining and Environmental Research group, Department of Statistics, University of Rajshahi, Rajshahi, Bangladesh; Department of Food Technology and Nutrition Science, Noakhali Science and Technology University, Noakhali-3814, Bangladesh

**Keywords:** COVID-19, Health Behavior, Principal Component Analysis (PCA), Socio-demographics, Bangladesh, Pakistan, United States

## Abstract

Control measures for the COVID-19 outbreak involve adequate public awareness and evaluation. These psychological and social aspects are effective in developing appropriate prevention and health communication strategies for mitigating the risk of an outcome. While a number of studies have investigated knowledge and awareness in response to the COVID-19 pandemic in specific populations, overall evaluation of these behavioral parameters in the context of different nations remained unexplored. This multinational comparative study is the first to investigate the COVID-19 knowledge, perception, and behavioral patterns among Bangladeshi, Pakistani, and United States individuals comprehensively, incorporating principal component analysis (PCA) along with traditional statistical analyses. Data from 646 participants of Bangladesh, Pakistan, and the United States were collected through an online questionnaire focusing on socio-demographics, information on knowledge, risk perception, precautionary measures, major information sources, and their trustworthiness. The findings revealed that participants from the United States were more knowledgeable (89.1%) than Pakistan and Bangladesh, respectively, whereas social media-based information was found to be a key predictor for lower knowledge levels (p = 0.04). Besides, females showed higher knowledge scores, while information from doctors and international health authorities positively influenced higher knowledge levels. Although the highest percentage of correct precautionary behavior was observed among the participants from the United States (85%), participants from Bangladesh showed a higher degree of worries. In addition to that, Pakistani people showed lower risk perception in response to this pandemic. Though further studies are needed to be conducted to address these circumstances, findings from this study might be helpful in implementing management policies, harm reduction, and risk communication protocols of the pandemic.

## 1 Introduction

COVID-19 emerged as a major global threat to human life several years ago. It has managed to spread globally to over 188 countries or regions, claiming more than 722,937 lives (Center for Systems Science and Engineering (CSSE) at Johns Hopkins University (JHU), n.d.). WHO immediately declared an emergency, implementing it in phases. IMST (Incident Management Support Team) was formed, a comprehensive technical package was declared, and strategic preparedness and response plans were circulated to fight the severity (WHO, 2020a). Despite reciprocated measures taken by the affected countries, confirmed cases and death tolls are mounting every day. Densely populated countries are particularly at higher risk during pandemics. Poverty and a poor healthcare system make the situation even dire. With 163 and 217 million people (UN DESAPD, 2019), two South Asian countries, Bangladesh and Pakistan, are the eighth and fifth most populous countries in the world, respectively. As lower middle-income economies (World Bank, 2020b), both countries lack the proper healthcare infrastructure, which limits public access to healthcare. In Bangladesh, per capita expenditure on healthcare is 36.3 USD, whereas in Pakistan it is 44.6 USD (World Bank, 2020a). Besides, the number of healthcare providers is insufficient; physicians per ten thousand people in Bangladesh and Pakistan are 5.3 and 9.8, respectively (WHO, 2020b). On the other hand, the United States is a high-income country with the highest healthcare expenditure per capita (World Bank, 2020a). Disease outbreak prevention relies on individual actions shaped by knowledge and risk perception. Informed individuals are more likely to adopt precautionary behaviors. Cultural practices can influence or override scientific knowledge, affecting responses. The source of information also has a major influence on knowledge and behavior during health crises (World Health Organization, 2020). Thus, the success of prevention relies on individual knowledge and action, supported by effective communication from healthcare professionals through trustworthy community channels.

Several studies reported the awareness and precautionary behaviors of specific populations against COVID-19. A cross-sectional study with the Bangladeshi population found a knowledge gap on COVID-19 transmission between healthcare professionals and the masses. The study also reported an age-related positive correlation to proper knowledge (Farhana & Mannan, 2020). Another study with educators (faculty and students) and healthcare workers (physicians, nurses, and lab staff) in Pakistan found inadequacy in preparation to stand against the ongoing pandemic (Khan et al., 2020). Yet other studies with physicians, pharmacists, and nurses in Pakistan have highlighted the importance of launching well-structured training programs for healthcare providers to improve their knowledge bases and better deal with the current situation (Hussain et al., 2020; Saqlain et al., 2020). Another group found that gender, education level, and area of residence were significantly associated with COVID-19-related knowledge (Hayat et al., 2020). A study conducted with the US population concluded that panic buying, attending large gatherings, and not wearing medical masks indicate lower COVID-19-related knowledge (Clements, 2020). Interestingly, political affiliation was found to be a determinant of knowledge in the same study. Other studies with Iranian (Taghrir et al., 2020), Chinese (Zhong et al., 2020), and Saudi Arabian (Alahdal et al., 2020) populations reported similar findings.

Although several studies reported COVID-19-related knowledge and awareness among a specific population, to our knowledge, a few comparative studies evaluated these behavioral parameters among residents of different nations. Thus, this research is an attempt to evaluate public knowledge, perceptions of risks and worries, precautionary behaviors, and trusted information sources related to COVID-19 in three countries—Bangladesh, Pakistan, and the United States. It also compared two lower-middle-income countries, Bangladesh and Pakistan, with a developed nation, the United States, in terms of individuals’ knowledge determinants and precautionary and diagnostic behaviors, as well as information sources.

The study examined the distribution of variables and characterized their distributions using frequencies with percentages. Comparisons between the groups were made by chi-square tests. A multivariate regression model was used to identify the factors associated with the outcome of interest. The knowledge scores and precautionary behavior scores of the study participants from three countries were considered the outcome variables. Initially, bivariate regression analyses were performed to identify the factors for multivariable analysis. Variables with P values <0.05 in the bivariate linear regression analysis were included in the multivariable models. In the multivariable linear regression models, associations were expressed as coefficients (β) with a standard error (SE), and a p-value of <0.05 was considered statistically significant. Principal component analysis (PCA) was performed to understand the question-wise (knowledge and precautionary behaviors) or category-wise (risk perception, worry and information source) variability across the three countries. These findings can inform targeted risk communication strategies by governments and health agencies, emphasizing the use of trusted information sources (like doctors and international health authorities) and counteracting misinformation from social media to improve public compliance and preparedness.

## 2 Methods

### 2.1 Study Area and Study Design

The current cross-sectional study is intended to assess the levels of knowledge, perception, and health behavior in response to the COVID-19 pandemic among social media users in three different countries—Bangladesh, Pakistan, and the United States. Bangladesh is in South Asia, between 20°34’ and 26°38’ N latitude and 88°01’ and 92°41’ E longitude, extending about 440 km east–west and 760 km north–south. Pakistan lies near 30° N and 70° E. The continental US spans 25°–50° N and 70°–125° W.

### 2.2 Data Collection Process

An electronic questionnaire was developed where the intention of the survey and assurance of the confidentiality of provided information, details about survey objectives and ethics and rights of the participants were briefly described, along with taking informed consent to proceed with participation in the study, which was clearly mentioned in the introductory section. Permanent residents between 18 and 49 years of age were the target participants. Four double-blinded sample collection campaigns were employed from April 30, 2020, to May 4, 2020, among 95,612 random Facebook users from selected countries. The survey appeared on news feeds, video feeds, instant articles and in stream videos of the audience network with no control from the researcher’s end over the distribution pattern. However, 751 Facebook users from the selected countries voluntarily participated in the survey. Through convenient sampling, researchers received 299 responses from Bangladesh, 216 from Pakistan, and 131 from the United States.

### 2.3 Measures

#### 2.3.1 Socio-demographic Factors

The socio-demographic data included information on age, sex, education, area of residence, employment status, and socio-economic status calculated based on their monthly income and educational status. However, socio-economic status was derived from their monthly income and was classified into five groups: lower income (monthly income less than 300 USD), lower middle income (monthly income ranges from 300 to 500 USD), higher middle income (monthly income ranges from 501 to 1000 USD), higher income (monthly income ranges from 1001 to 2000 USD), and upper income (monthly income more than 2000 USD). Besides, employment status was classified into three groups: unemployed (students and those who were not enrolled in any kind of paid job), self-employed (freelancing and business), or involved with other self-earnings.

#### 2.3.2 Knowledge assessment

Nine basic COVID-19-related statements (Table 2) estimated the knowledge level. Considering an incorrect response as ‘0’ and a correct response as ‘1’, a sum score (theoretical range 0-9) was calculated and used for descriptive analysis. Overall knowledge scores were also used for linear regression analysis.

#### 2.3.3 Worries and risk perception assessment

To assess risk perception, the participants were asked to rank their own risk, the perceived risk of their close relations and their country of residence in either the low, intermediate, or high categories. Worries about proper treatment, consumables and medical supplies were also assessed on the same scale. The descriptive analysis maintained the same categories for both risk perception and worry analyses.

#### 2.3.4 Precautionary behaviour assessment

Precautionary behaviour was evaluated by 7 questions (Table 5), answers to which were enumerated (not cautious=0, cautious=1) to get a sum score (theoretical range 0-7). Overall scores for precautionary behaviours (theoretical range: 0-7) were used for descriptive and linear regression analysis. Sources of information and their reliability were also enumerated based on the responses (ticked=1, not ticked=0). The number of responses against each source was calculated for the descriptive and linear regression analysis.

### 2.4 Statistical Analysis

#### 2.4.1 Chi-Square (χ^2^) test

The Chi-Square (χ^2^) test is a non-parametric statistical method used to evaluate the relationship or independence between two categorical variables, where the data from both variables is represented as values. Chi-Square is also called Kai Squared. It evaluates whether the observed frequency distribution of data differs significantly from the expected distribution under the null hypothesis of no association (Franke et al., 2012). The Chi-Square test uses a cross-tabulation table to compare observed and expected frequencies, helping to test the independence between categorical variables (Pandis, 2016). The test statistic is calculated as:

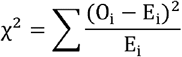

where O_i_ and E_i_ represent the observed and expected frequencies for each category, respectively. The significance of the test depends upon the comparison of the computed χ^2^ with a critical value for the Chi-Square distribution table for a selected confidence level (commonly, α = 0.05). The degrees of freedom for the test will rely on the number of categories for each of the variables.

#### 2.4.2 Principal Component Analysis (PCA)

Principal Component Analysis (PCA) is a multivariate statistical technique used to lower dimensions so as to preserve as much variability as possible in the data (Abdi & Williams, 2010). In this study, it was used to determine the underlying structure and variability in responses with the respect to knowledge, precautionary behavior, risk perception, concern, and information towards the three countries. To perform PCA, the data matrix X was first standardized. The covariance matrix C was computed as:

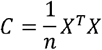

PCA involves solving the eigenvalue decomposition of the covariance matrix:

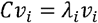

Where:λ*_i_* is the eigenvalue representing the variance explained by the i-th component *v_i_*, is the corresponding eigenvector (loading), and C is the covariance matrix of the standardized variables.

#### 2.4.3 Multivariate factors

The multivariate factors were arranged in the X matrix and reduced into a product of two new matrices by using the following equation.

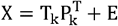

Here, T_k_ is the matrix of scores, which signifies the relation of sample to each other, P_k_ is the matrix of loadings, carrying information about the relation of variables to each other, k is the number of factors into the model and E is the unmodeled variance.

All the statistical analyses were performed using R version 4.4.2 software (https://www.r-project.org), Foundation for Statistical Computing).

## 3 Results

### 3.1 Characteristics of the participants

Overall, 646 participants were included in this analysis. Among them, 299, 216, and 131 were from Bangladesh, Pakistan, and the United States, respectively. Significant differences were observed in background demographic characteristics across the countries. However, more than two-thirds of the Pakistani participants were male (80.1%) and about two-thirds of the Bangladeshi participants were male (65.5%). In addition to that, more female participation was from the United States (67.4%).

Young adults (18–29) dominated in Bangladesh (73.9%) but were lower in the U.S. (45.8%), where older respondents (40–49: 24.4%) were more frequent. Pakistan had more high school or below (14.4%) education as compared to U.S. more graduate or above (35.1%). Bangladesh had the highest rate of unemployment (53.5%) and the U.S. had the highest rate of employment (51.9%). Incomes differed significantly: under 300 USD in Bangladesh (57.9%) and Pakistan (63.9%) and over 2000 USD in the U.S (34.4%) **(Table 1)**.

**Table 1.** Background characteristics of the respondents.

| Variables | Countries |  |  | Chi-square Test Statistics |  |
| --- | --- | --- | --- | --- | --- |
| | Bangladesh | Pakistan | United States | $\chi^2$ test value | p-value |
| Gender |  |  |  |  |  |
| Male | 193; 65.5% | 173; 80.1% | 44; 32.6% | 78.95 | <0.001 |
| Female | 106; 34.5% | 43; 19.9% | 87; 67.4% |  |  |
| Age |  |  |  |  |  |
| Between 18-29 | 221; 73.9% | 151; 69.9% | 60; 45.8% | 78.88 | <0.001 |
| Between 30-39 | 75; 25.1% | 51; 23.6% | 39; 29.8% |  |  |
| Between 40-49 | 3; 1% | 14; 6.5% | 32; 24.4% |  |  |
| Educational status |  |  |  |  |  |
| High school or below | 15; 5% | 31; 14.4% | 28; 21.4% | 39.92 | <0.001 |
| Undergraduate | 84; 28.1% | 55; 25.5% | 37; 28.2% |  |  |
| Graduate or above | 111; 37.1% | 67; 31% | 46; 35.1% |  |  |
| Employment status |  |  |  |  |  |
| Unemployed | 160; 53.5% | 94; 43.5% | 46; 35.1% | 20.53 | <0.001 |
| Self-employed | 25; 8.4% | 38; 17.6% | 17; 13% |  |  |
| Employed | 114; 38.1% | 84; 38.9% | 68; 51.9% |  |  |
| Monthly income |  |  |  |  |  |
| <300 USD | 173; 57.9% | 138; 63.9% | 37; 28.2% | 176.63 | <0.001 |
| 300-500 USD | 64; 21.4% | 39; 18.1% | 9; 6.9% |  |  |
| 501-1000 USD | 34; 11.4% | 26; 12% | 12; 9.2% |  |  |
| 1001-2000 USD | 18; 6% | 7; 3.2% | 28; 21.4% |  |  |
| >2000 USD | 10; 3.3% | 6; 2.8% | 45; 34.4% |  |  |

### 3.2 Knowledge level

Overall, participants from the United States were found more knowledgeable about COVID-19 (89.1% correct answers) than those of Bangladesh (85.4%) and Pakistan (75.2%). However, no significant difference across the countries was found to be aware of COVID-19 transmission from pets (χ^2^=4.66, p=0.097). On the other hand, statement-wise correct response percentages observed low among Pakistani participants (75.2%) compared to the other two nations. Regarding the risk of false results from RT-PCR-based diagnosis, people from both Bangladesh (61.3%) and Pakistan (53.3%) had significantly lower-level knowledge than the United States (81.5%) **(Table 2)**.

**Table 2.** Level of Knowledge on COVID-19 among the participants.

| Statements<br>(Yes or No) | Correct Answer Rate (%) |  |  | Chi-square Test Statistics |  |
| --- | --- | --- | --- | --- | --- |
| | Bangladesh | Pakistan | United States | $\chi^2$ test value | p-value |
| Novel Corona-virus (SARS-CoV-2) cannot be transmitted in hot and humid climates. (N) | 91.5% | 79.6% | 87.8% | 15.51 | <0.001 |
| Novel Corona-virus can survive on the outer surface of surgical masks. (Y) | 93.3% | 85.1% | 95.4% | 13.95 | <0.001 |
| Patients with COVID-19 may manifest symptoms like diarrhoea, nausea, or headache. (Y) | 85.2% | 63.3% | 89.6% | 48.06 | <0.001 |
| People with no symptoms can transmit the virus. (Y) | 94.6% | 86.5% | 96.9% | 16.44 | <0.001 |
| Mild symptoms can be managed at home. (Y) | 93.3% | 87.5% | 96.2% | 9.66 | 0.008 |
| COVID-19 can be transmitted from pets. (N) | 61.1% | 56.3% | 67.9% | 4.66 | 0.097 |
| Once recovered, the patients are at no risk of re-infection. (N) | 90.6% | 76.9% | 90.8% | 22.7 | <0.001 |
| Elderly population are at higher risk of mortality. (Y) | 94.9% | 91.2% | 99.2% | 10.48 | 0.005 |
| Corona-virus testing using RT-PCR for COVID-19 detection can provide | 61.3% | 53.3% | 81.5% | 28.00 | <0.001 |
| false result. (Y) |  |  |  |  |  |
| **Total** | **85.4%** | **75.2%** | **89.1%** |  |  |

### 3.3 Precautionary measures

Table 3 illustrated precautionary measures taken in response to COVID-19 among the participants across the countries. In spite of having a high level of knowledge, participants from the United States were found to manifest less caution (74.6% correct behavior) than that of Pakistan (76.2%) or Bangladesh (87.4%) adjacently. Besides, question-wise differences were significant across the countries except in the answers related to home deliveries (χ^2^=0.17, p=0.092) and hand washing (χ^2^=0.90, p=0.64). However, visits to the nearby shops or local markets were more frequent in Pakistani participants (49.8%). On the other hand, a lower cautiousness of home delivery of restaurant foods was observed in US participants (45.8%) **(Table 3)**.

**Table 3.**
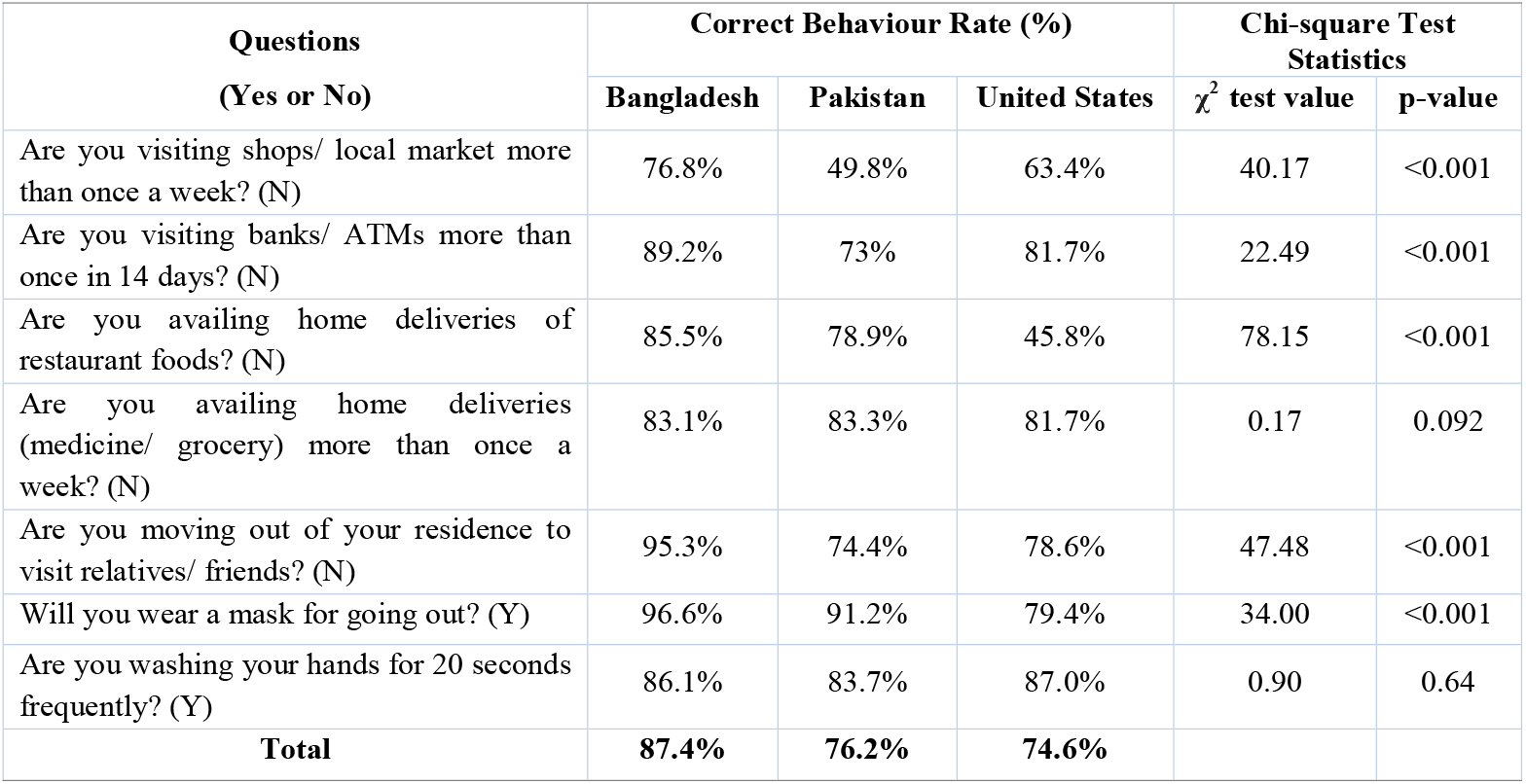
Precautionary measures against COVID-19 among the participants.

| Questions<br>(Yes or No) | Correct Behaviour Rate (%) |  |  | Chi-square Test Statistics |  |
| --- | --- | --- | --- | --- | --- |
| | Bangladesh | Pakistan | United States | $\chi^2$ test value | p-value |
| Are you visiting shops/ local market more than once a week? (N) | 76.8% | 49.8% | 63.4% | 40.17 | <0.001 |
| Are you visiting banks/ ATMs more than once in 14 days? (N) | 89.2% | 73% | 81.7% | 22.49 | <0.001 |
| Are you availing home deliveries of restaurant foods? (N) | 85.5% | 78.9% | 45.8% | 78.15 | <0.001 |
| Are you availing home deliveries (medicine/ grocery) more than once a week? (N) | 83.1% | 83.3% | 81.7% | 0.17 | 0.092 |
| Are you moving out of your residence to visit relatives/ friends? (N) | 95.3% | 74.4% | 78.6% | 47.48 | <0.001 |
| Will you wear a mask for going out? (Y) | 96.6% | 91.2% | 79.4% | 34.00 | <0.001 |
| Are you washing your hands for 20 seconds frequently? (Y) | 86.1% | 83.7% | 87.0% | 0.90 | 0.64 |
| **Total** | **87.4%** | **76.2%** | **74.6%** |  |  |

### 3.4 Risk perception and worry

The countries also varied significantly in terms of individual, familial or country risk, whereas a higher proportion of Pakistani subjects considered them or their families to be at intermediate or lower risk during the pandemic. On the other hand, responders from the United States (68.7%) and especially Bangladesh (86.2%) considered their countries to be at higher risk. However, participants from Bangladesh showed more worry about appropriate treatment, consumable supply, and medical supplies than those of the other two nations. For the other two countries, the worry was shared closely in the three different levels among the three categories **(Figure 2)**.

**Figure 1.**
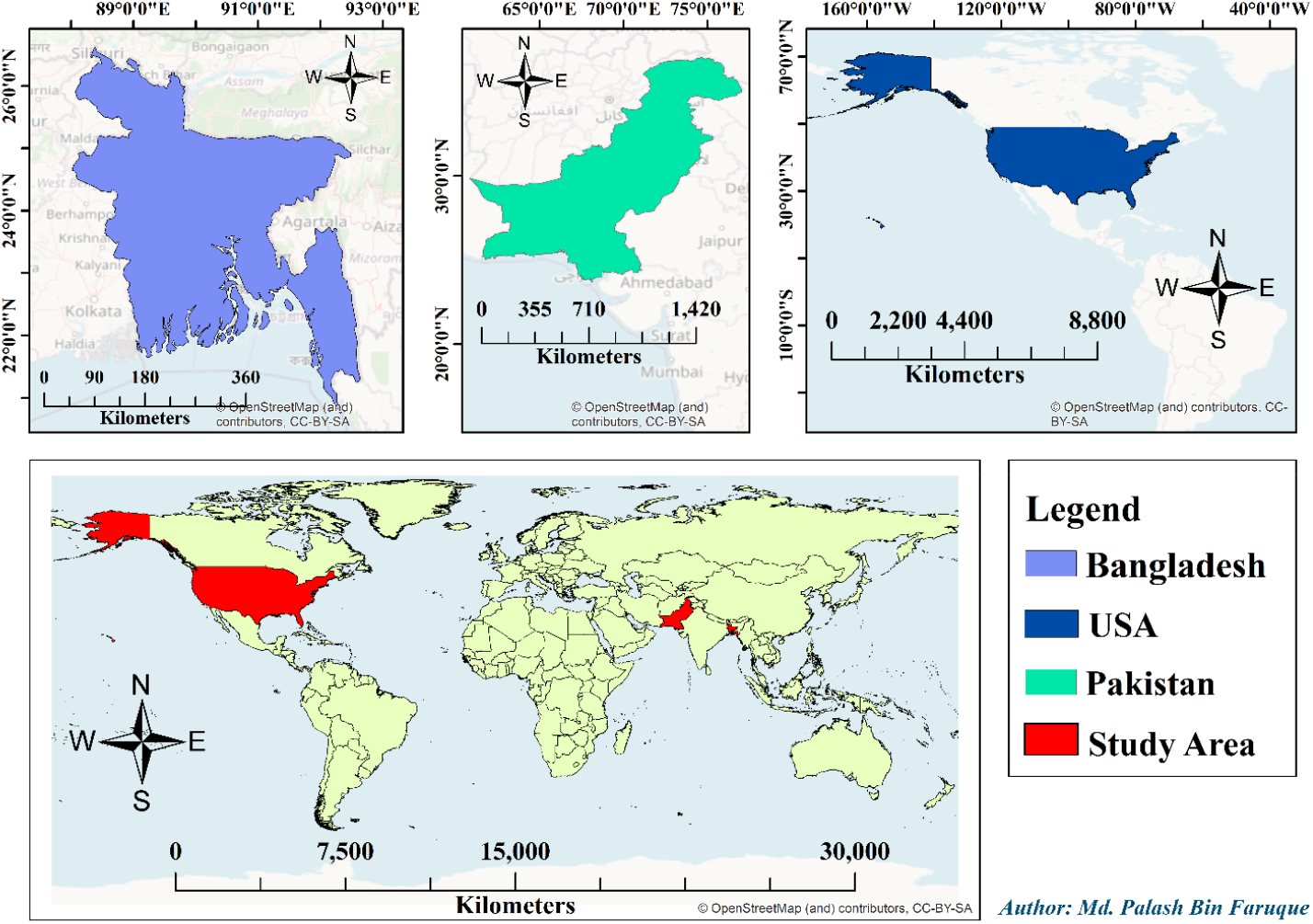
Geographical representation of the study areas including regional maps of the United States, Pakistan, and Bangladesh, overlaid with a reference to their positions on the world map Using extensive literature review, a questionnaire was made which was piloted with about 20 residents of Bangladesh. The online survey comprised five sections: (i) level of COVID-19-related knowledge, (ii) degree of risk perception, (iii) worries about food and medical supplies, (iv) behavioral and precautionary attitude, and (v) COVID-19-related information sources. However, a non-probability sampling technique (convenience sampling) was employed for this study; data were gathered via an online questionnaire distributed through Facebook.

**Figure 2.**
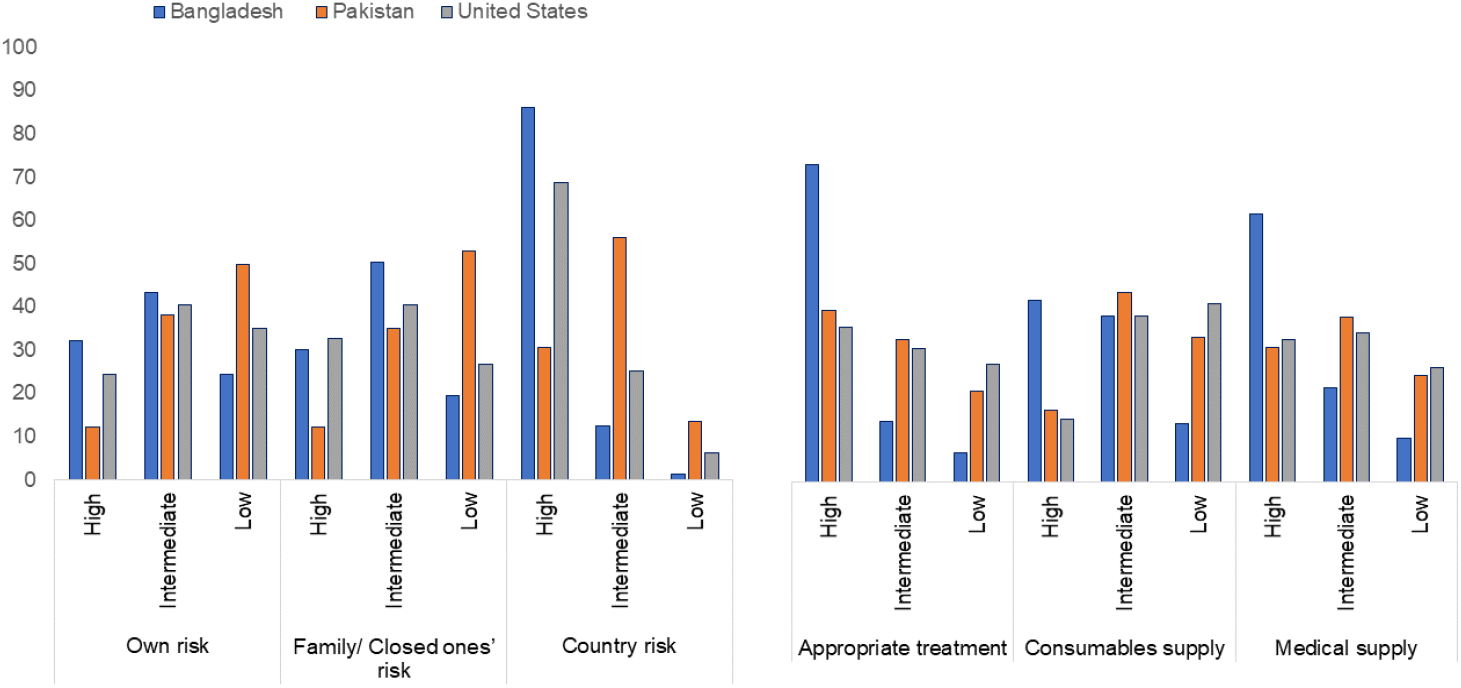
Level of COVID-19 related risk perception and worry in the study participants.

### 3.5 Information Source

Social media accounted for the most common source of COVID-19 information in nations (89.6% Bangladesh, 83.6% Pakistan, 75.4% USA). Government (55–69%) and newspaper sources (21–46%) were used moderately, with doctors (36–37%) and friends (28–47%) to a lesser extent, showing population heterogeneity in reliance on information sources. Besides, significant differences were observed in the use of newspapers (χ^2^=33.17, *p* =<0.001), social media (χ^2^=14.32, *p* =<0.001), and friends (χ^*2*^*=20*.*37, p =<0*.*001*) as the major information sources across the three countries. Dependability of newspaper (χ^2^=25.82, *p*=<0.001), television (χ^2^=14.78, *p*=<0.001), government sources (χ^2^=20.02, *p*=<0.001), doctors (χ^2^=26.34, *p*=<0.001), and international health authorities (χ^2^=14.90, *p*=<0.001) as information sources also differed significantly among the countries. Apart from that, a higher proportion of study participants from Bangladesh (37.2%) regarded newspapers as a more reliable source than those of Pakistan (17.5%) or the United States (22.8%), although it showed poor overall trustworthiness (28%).

Moreover, only 41.2% of Bangladeshi participants expressed more trust in government sources than that of Pakistan (60.4%) or the United States (56.1%). Participants from the United States were considering physicians as a reliable information source more than Bangladeshis or Pakistanis **(Table 4)**.

**Table 4.** Major sources of COVID-19 related information and their reliability among the participants.

| Information Sources | Major Sources |  |  |  |  | Reliability |  |  |  |  |
| --- | --- | --- | --- | --- | --- | --- | --- | --- | --- | --- |
|  | Bangladesh (n;%) | Pakistan (n;%) | United States (n;%) | Chi-square Test Statistics |  | Bangladesh (n;%) | Pakistan (n;%) | United States (n;%) | Chi-square Test Statistics |  |
| | | | | $\chi^2$ test value | p-value | | | | $\chi^2$ test value | p-value |
| Newspaper |  |  |  |  |  |  |  |  |  |  |
| Yes | 135; 45.5% | 45; 21% | 42; 32.3% | 33.17 | <0.001 | 110; 37.2% | 37; 17.5% | 28; 22.8% | 25.82 | <0.001 |
| No | 162; 54.5% | 169; 79% | 88; 67.7% |  |  | 186; 62.8% | 175; 82.5% | 95; 77.2% |  |  |
| Social media |  |  |  |  |  |  |  |  |  |  |
| Yes | 266; 89.6% | 179; 83.6% | 98; 75.4% | 14.32 | <0.001 | 70; 23.6% | 56; 26.4% | 23; 18.7% | 2.57 | 0.28 |
| No | 31; 10.4% | 35; 16.4% | 32; 24.6% |  |  | 226; 76.4% | 156; 73.6% | 100; 81.3% |  |  |
| Television |  |  |  |  |  |  |  |  |  |  |
| Yes | 192; 64.6% | 127; 59.3% | 78; 60% | 1.74 | 0.42 | 133; 44.9% | 65; 30.7% | 36; 29.3% | 14.78 | <0.001 |
| No | 105; 35.4% | 87; 40.7% | 52; 40% |  |  | 163; 55.1% | 147; 69.3% | 87; 70.7% |  |  |
| Government Source |  |  |  |  |  |  |  |  |  |  |
| Yes | 163; 54.9% | 131; 61.2% | 90; 69.2% | 7.98 | 0.018 | 122; 41.2% | 128; 60.4% | 69; 56.1% | 20.02 | <0.001 |
| No | 134; 45.1% | 83; 38.8% | 40; 30.8% |  |  | 174; 58.8% | 84; 39.6% | 54; 43.9% |  |  |
| Doctors |  |  |  |  |  |  |  |  |  |  |
| Yes | 108; 36.4% | 64; 29.9% | 48; 36.9% | 2.79 | 0.25 | 129; 43.6% | 82; 38.7% | 82; 66.7% | 26.34 | <0.001 |
| No | 189; 63.6% | 150; 70.1% | 82; 63.1% |  |  | 167; 56.4% | 130; 61.3% | 41; 33.3% |  |  |
| Friends |  |  |  |  |  |  |  |  |  |  |
| Yes | 142; 47.8% | 60; 28% | 53; 40.8% | 20.37 | <0.001 | 26; 8.8% | 16; 7.5% | 11; 8.9% | 0.30 | 0.86 |
| No | 155; 52.2% | 154; 72% | 77; 59.2% |  |  | 270; 91.2% | 196; 92.5% | 112; 91.1% |  |  |
| International health authorities |  |  |  |  |  |  |  |  |  |  |
| Yes | 186;<br>62.6% | 114;<br>53.3% | 89;<br>68.5% | 8.69 | 0.013 | 241;<br>81.4% | 141;<br>66.5% | 94;<br>76.4% | 14.90 | <0.001 |
| No | 111;<br>37.4% | 100;<br>46.7% | 41;<br>31.5% |  |  | 55;<br>18.6% | 71;<br>33.5% | 29;<br>23.6% |  |  |

**Table 5.** Multivariable linear regression analysis to assess the factors associated with knowledge scores among the study participants across three countries.

| Variables | Unadjusted model |  | Adjusted model |  |
| --- | --- | --- | --- | --- |
| | $\beta$ (SE) | p-value | $\beta$ (SE) | p-value |
| **Age** | 0.03 (0.01) | <0.001 | 0.02 (0.01) | 0.09 |
| **Gender** |  |  |  |  |
| Male | -0.54 (0.11) | <0.001 | -0.31 (0.12) | 0.007 |
| Female | Reference category |  |  |  |
| **Country of residence** |  |  |  |  |
| Bangladesh | -0.40 (0.14) | 0.003 | -0.20 (0.17) | 0.24 |
| Pakistan | -1.25 (0.14) | <0.001 | -0.79 (0.18) | <0.001 |
| United States | Reference category |  |  |  |
| **Educational status** |  |  |  |  |
| High school or below | -0.43 (0.17) | 0.01 | -0.25 (0.17) | 0.14 |
| Undergraduate | -0.33 (0.12) | 0.008 | -0.18 (0.13) | 0.17 |
| Graduate or above | Reference category |  |  |  |
| **Monthly income** |  |  |  |  |
| <300 USD | -0.83 (0.19) | <0.001 | -0.27 (0.21) | 0.20 |
| 300-500 USD | -0.50 (0.21) | 0.02 | -0.06 (0.23) | 0.81 |
| 501-1000 USD | -0.34 (0.24) | 0.15 | 0.01 (0.24) | 0.97 |
| 1001-2000 USD | -0.04 (0.25) | 0.87 | 0.09 (0.24) | 0.71 |
| >2000 USD | Reference category |  |  |  |
| **Confidence on Doctors (Yes)** | 0.47 (0.11) | <0.001 | 0.25 (0.11) | 0.02 |
| **Confidence on Newspapers (Yes)** | 0.29 (0.12) | 0.02 | 0.19 (0.11) | 0.10 |
| **Confidence on social media (Yes)** | -0.50 (0.13) | <0.001 | -0.26 (0.12) | 0.04 |
| **Confidence on International Authorities (Yes)** | 0.51 (0.13) | <0.001 | 0.30 (0.12) | 0.02 |
| **Total Behavioural Score** | 0.13 (0.04) | 0.002 | 0.07 (0.04) | 0.10 |

### 3.6 Principal component analysis

The countries behaved differently, as their relative positions and clustering patterns differed in principal component analysis. In terms of overall response, a fair similarity was observed between the participants from Pakistan and the United States. Question-wise knowledge and category-wise risk perception differed the most in participants from Bangladesh and the least from the United States. Besides, risk perception and knowledge were found to be highly correlated, whereas information source and precautionary behaviors showed a strong association. Moreover, knowledge and worry were more correlated than precautionary measures and worry **(Figure 3)**.

**Figure 2.**
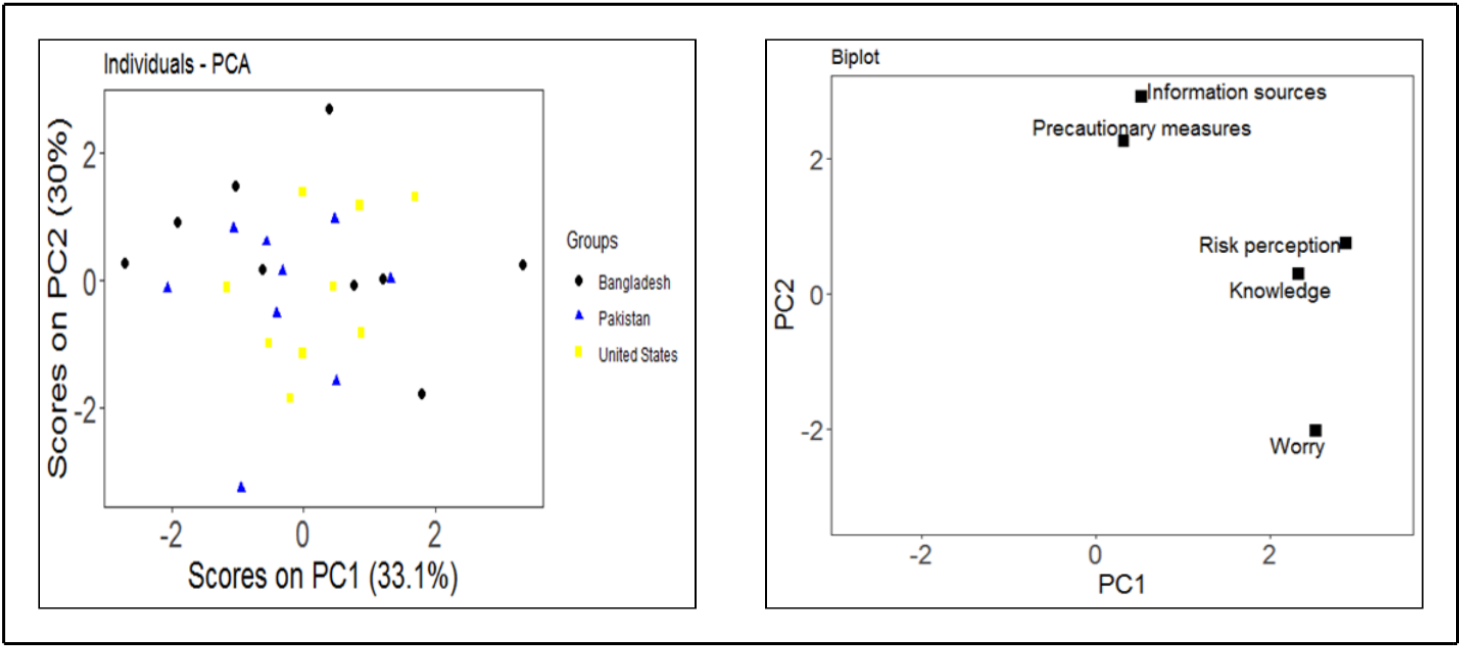
Scores and loading plots of the three study groups.

### 3.7 Multivariable Linear Regression Analysis

Multivariate linear regression analysis for knowledge scores suggested male participants to be less knowledgeable (adjusted β = −0.31, p = 0.007) than females. Besides, a significantly lower level of knowledge was found in Pakistani participants (adjusted β = −0.79, p < 0.001) than that of the US, whereas the knowledge difference between Bangladeshi and US samples came out as non-significant. In terms of information source, having confidence in social media-based information was a determinant of lowering knowledge (adjusted β = −0.26, p = 0.04), whereas confidence in doctors (adjusted β = 0.25, p = 0.02) and the international health authorities (adjusted β = 0.30, p = 0.02) showed positive correlation with higher knowledge scores **(Table 5)**.

In the multivariate linear regression analysis for precautionary behaviors, female participants were found to be noticeably less cautious (adjusted β = −0.45, *p* < 0.001) than males. However, Bangladeshi participants maintained a significantly higher degree of caution (adjusted β = 0.90, *p* < 0.001) than that of US participants. Besides, participants who were self-employed (β *= −0*.*38, p =* 0.03) were more cautious than unemployed and employed participants, respectively. Besides, regarding the participant’s information source, where confidence in social media-based information (adjusted β = −0.35, *p =* 0.003) was two key determinants of lower precautions. To a nearly significant extent, as predicted, a higher knowledge score influenced the precautionary behavior positively (adjusted β = 0.06, p = 0.09) **(Table 6)**.

**Table 6.** Multivariable linear regression analysis to assess the factors associated with precautionary behaviors scores among the study participants across three countries.

| Variables | Unadjusted model |  | Adjusted model |  |
| --- | --- | --- | --- | --- |
| | $\beta$ (SE) | p-value | $\beta$ (SE) | p-value |
| Age | -0.03 (0.01) | <0.001 | -0.01 (0.01) | 0.38 |
| Gender |  |  |  |  |
| Female | -0.38 (0.11) | <0.001 | -0.45 (0.11) | <0.001 |
| Male | Reference category |  |  |  |
| Country of residence |  |  |  |  |
| Bangladesh | 0.94 (0.13) | <0.001 | 0.90 (0.16) | <0.001 |
| Pakistan | 0.16 (0.14) | 0.25 | 0.32 (0.17) | 0.06 |
| United States | Reference category |  |  |  |
| Education |  |  |  |  |
| High school or below | -0.30 (0.17) | 0.09 | -0.08 (0.17) | 0.64 |
| Undergraduate | 0.24 (0.13) | 0.06 | 0.10 (0.14) | 0.46 |
| Graduate or above | Reference Category |  |  |  |
| Occupation |  |  |  |  |
| Employed | -0.36 (0.11) | <0.001 | -0.13 (0.15) | 0.39 |
| Self-employed | -0.73 (0.16) | <0.001 | -0.38 (0.18) | 0.03 |
| Unemployed | Reference Category |  |  |  |
| Monthly income |  |  |  |  |
| <300 USD | 0.67 (0.18) | <0.001 | 0.15 (0.21) | 0.46 |
| 300-500 USD | 0.56 (0.21) | 0.006 | 0.07 (0.22) | 0.75 |
| 501-1000 USD | 0.51 (0.23) | 0.02 | 0.08 (0.23) | 0.73 |
| 1001-2000 USD | 0.47 (0.24) | 0.05 | 0.22 (0.23) | 0.34 |
| >2000 USD | Reference Category |  |  |  |
| Confidence on social media (Yes) | -0.39 (0.12) | 0.001 | -0.35 (0.12) | 0.003 |
| Confidence on International Authorities (Yes) | 0.37 (0.12) | 0.002 | 0.15 (0.12) | 0.19 |
| Total knowledge Score | 0.12 (0.04) | 0.002 | 0.06 (0.04) | 0.09 |

## 4 Discussion

This study investigates the level of knowledge, risk perception, worries, precautionary behavior, and major COVID-19-related information sources of the participants from Bangladesh, Pakistan, and the United States. Besides, Bangladesh and Pakistan were evaluated on the above-mentioned categories relative to the United States. A deeper look into the variables also revealed the significant determinants of the COVID-19-related knowledge and precautionary behaviors against the outbreak.

This study observed an overall high level of COVID-19-related knowledge in the participants, which goes in parallel with the findings of several other groups (Taghrir et al., 2020; Zhong et al., 2020). Participants from the US were found to be more well-informed. As females were higher in proportion in the US samples, females are concluded to possess more knowledge regarding COVID-19. However, studies suggest the gender-specific degree of knowledge to be elusive, as some found females to be more knowledgeable (Azlan et al., 2020; Clements, 2020; Hussain et al., 2020; Zhong et al., 2020), whereas few others concluded the opposite (Alahdal et al., 2020; Hayat et al., 2020). The least awareness of COVID-19 transmission from pets across the participants could be due to various news portals, which speculated about the transmission from pets being able to transmit the virus to another human being (Parry, 2020). Although lately, health authorities maintained a rare possibility of spreading (CDC, 2020), during the time of response collection, authorities including WHO were against the plausibility of pet-to-human transmission. However, as a result of continuous education programmes and regular clinical practices, physicians are presumed to be more knowledgeable than the others. Thus, it was justified to observe that relying on physicians for COVID-19-related information was a significant predictor of a higher knowledge level. On the other hand, relying on information from social media can be inaccurate or harmful. Gimmick information, false news, and exaggerated claims flood social media. Unsubstantiated information circulated on social media is often accepted by the common people naively due to the providers’ eloquence or educational background. The participants from the US regarded social media as their least preferred source of COVID-19-related information and were justified as being more knowledgeable than the participants from the other two countries. While social media is regarded both as a blessing and a curse (theconversation.com, 2020), WHO is leading the fight against the ‘infodemic’ through working with social media giants (www.scmp.com, 2020). Given the up-to-date information they are receiving, it came out as no surprise to find people who put confidence in international health authorities to be more knowledgeable.

Studies with the participants from Hubei, China (Zhong et al., 2020); Iran (Taghrir et al., 2020); Malaysia (Azlan et al., 2020); and Saudi Arabia (Alahdal et al., 2020) concluded that the people from those areas showed proper precautionary behaviors. However, studies on Pakistan (Khan et al., 2020; Saqlain et al., 2020), Bangladesh (Farhana & Mannan, 2020), and US (Clements, 2020) population found inadequate precautionary practices. In this study, US subjects were found to be less cautious, probably due to the lack of coordination by the US government to impose strict lockdown measures. Clement suggests the same in his study (Clements, 2020). Besides, the precaution regarding availing the deliveries from restaurants is an important factor to be mentioned. Since the US is the second largest online food delivery market across the globe (www.statista.com, 2020), people in the US are ahead of countries like Bangladesh or Pakistan in terms of food delivery trends. Thus, their seemingly normal behavioral pattern was regarded as less cautious during the pandemic and influenced the overall precautionary behavior. Disregarding this factor might support the findings of Clement, who observed that an increase in knowledge scores improved precautionary behavior (Clements, 2020).

Participants from Pakistan may have a lower risk perception due to their limited knowledge about COVID-19. Participants from Bangladesh considered their country to be at high risk, although perceived self- or familial risks were lower in the same group. Multiple factors could explain this disparity. Since the detection of the first corona patient, shortcomings were observed in preparedness and response. Point of entry control and contact tracing were questionable, which mass media addressed oftentimes. Healthcare facilities in Bangladesh were dire; the number of hospital beds, ICU facilities, and medical equipment was low, becoming a big concern. Protective equipment for doctors was sparse, and there were loopholes in the imposed lockdown and travel restrictions. The observations made by the UNDP (United Nations Development Program) were consistent with these concerns (UNDP, 2020). Although people preferred staying home and ensured personal and family safety, their perceived risk about the country was higher, which was reflected in the study. On the other hand, the participants from Bangladesh were more worried about appropriate treatment, consumables, and medical supplies than those of Pakistan and the United States. The observed higher worry can be explained by considering the scarcity of hand gloves, masks, sanitizers, and immuno-enhancer medicines like zinc sulphate or ascorbic acid. The worries regarding treatment in Bangladesh were also addressed by UNDP (UNDP, 2020).

Social media was the most common information source (89.6%) for both Bangladeshi (89.6%) and Pakistani (83.6%) subjects, which is supported by other studies (Farhana & Mannan, 2020; Saqlain et al., 2020). As confidence on social media was indicative of lower COVID-19-related knowledge, it must be kept in high consideration while taking any initiative to disseminate correct information. Since Bangladeshis demonstrated confidence in international health authorities, dissemination of the communication material of the global health authorities will make a positive change. Furthermore, government initiatives will lessen worries and perceived risks. Subjects from Pakistan demonstrated the lowest level of knowledge among all. Khan et al. also reported the inadequacy of knowledge among front-line healthcare workers (Khan et al., 2020). Although healthcare professionals were found highly cautious (Hussain et al., 2020; Saqlain et al., 2020), this study with a random sample from Pakistan showed it to be noticeably lower. Higher confidence in the governmental information sources in Pakistan will help raise awareness by making information from doctors and health authorities more available and popular. In the US subjects, confidence in doctors and international health authorities positively influenced the knowledge level, which is consistent with other study findings (Clements, 2020). Other studies with US populations suggest that the behavioral patterns of US citizens are highly influenced by governmental directions on pandemic management (Clements, 2020; Czeisler et al., 2020). While stay-at-home orders and business closures are supported by the majority, a high degree of adherence to COVID-19 mitigation guidelines is also there (Czeisler et al., 2020). Since the people are already knowledgeable and welcoming towards government direction, countrywide coordination of mitigation is pivotal.

The sampling conducted through an electronic questionnaire on social media exhibited some bias. Underprivileged or illiterate people, especially from Bangladesh and Pakistan, might have been overlooked due to their narrow exposure to online platforms. Participation of elder people was also less due to their unfamiliarity with social media. Yet, considering the stay-at-home orders prevailing at that time in all three countries, an online questionnaire was the best way to collect responses.

## 5. Conclusion

This study found country-wise differences in COVID-19-related knowledge, risk perception, and precautionary behavior. Females were more knowledgeable, while confidence in information from doctors and global health authorities was found to significantly raise the knowledge level. Social media, despite being a major information source, provided information that was associated with low knowledge levels and incorrect precautionary behavior. People from Bangladesh were found to be more worried, while lower risk perception in Pakistani people is associated with their low knowledge level. Despite having a high knowledge level, people from the United States were maintaining low precautionary measures. Awareness programs in light of global health authorities and through the doctor community are essential with guidance about correct information sources. While further studies regarding public awareness and health behavior are important to conduct, the findings from this study might be helpful in the planning and implementation of pandemic management policies for the respective countries.

## Data Availability

All data produced in the present study are available upon reasonable request to the authors

## 6 Declaration

### 6.1 Ethical approval and consent to participate

Institutional Animal, Medical Ethics, Biosafety and Biosecurity Committee (IAMEBBC) for Experimentations on Animal, Human, Microbes and Living Natural Sources at the Institute of Biological Sciences, University of Rajshahi approved the study. This study was conducted in full compliance with research ethics norms and, more specifically, the scientific principles of human research ethics established in the Declaration of Helsinki. It was also conducted following the Checklist for Reporting Results of Internet E-Surveys (CHERRIES) guidelines. The participants were informed of the purpose and objectives of the study through a brief description at the start of the questionnaire, and they were asked about their consent to their participation in it. They were assured that they had the freedom to quit the provided lint at any point in time. No monetary or non-monetary remuneration or benefit was offered for participating in the study.

### 6.2 Availability of data and materials

The datasets will be made available to appropriate academic parties upon request from the corresponding author.

### 6.5 Author’s contribution

This study was initially conceptualized by all the team members. Md. Abdur Rahman and Saiful Islam conceived the study and developed the methodology with Md. Shakibul Hasan. Saiful Islam has managed software tasks. Md. Abdul Khalek and Md. Mostafizur Rahman validated the results. Md. Abdur Rahman, Saiful Islam and Md. Palash Bin Faruque handled analysis, data curation, and visualization. The investigation involved authors Md. Abdur Rahman, Saiful Islam, Md. Palash Bin Faruque and Md. Shakibul Hasan, with resources provided by Md. Shakibul Hasan. Md. Abdur Rahman and Saiful Islam drafted the manuscript; Md. Mostafizur Rahman and Md. Abdul Khalek reviewed and edited it. Md. Mostafizur Rahman supervised the project, which was administered by Md. Abdur Rahman. The project was implemented by NBM, M. Akther, TM, ABS, M. Akter, MS. Ahmed, MSMS, AAH, and MAM; they participated in the data collection and management which was administered by Mst. Shabrina Afroz. The study area map made by the Md. Palash Bin Faruque. All authors approved the final manuscript.

